# Indirect Impact of the COVID-19 Pandemic on Activity and Outcomes of Transcatheter and Surgical Treatment of Severe Aortic Stenosis in England

**DOI:** 10.1101/2020.08.05.20168922

**Authors:** Glen P. Martin, Nick Curzen, Andrew T. Goodwin, James Nolan, Lognathen Balacumaraswami, Peter Ludman, Evangelos Kontopantelis, Jianhua Wu, Chris Gale, Mark A de Belder, Mamas A. Mamas

## Abstract

**Background:** Aortic stenosis requires timely treatment with either surgical aortic valve replacement (SAVR) or transcatheter aortic valve replacement (TAVR). This study aimed to investigate the indirect impact of COVID-19 on national SAVR and TAVR activity and outcomes.

**Methods:** The UK TAVR Registry and the National Adult Cardiac Surgery Audit were used to identify all TAVR and SAVR procedures in England, between January 2017 and June 2020. The number of isolated AVR, AVR+coronary artery bypass graft (CABG) surgery, AVR+other surgery and TAVR procedures per month was calculated. Separate negative binomial regression models were fit to monthly procedural counts, with functions of time as covariates, to estimate the expected change in activity during COVID-19.

**Results:** We included 13376 TAVR cases, 12328 isolated AVR cases, 7829 AVR+CABG cases, and 6014 AVR+Other cases. Prior to March 2020 (UK lockdown), monthly TAVR activity was rising, with a slight decrease in SAVR activity during 2019. We observed a rapid and significant drop in TAVR and SAVR activity during the COVID-19 pandemic, especially for elective cases. Cumulatively, over the period March to June 2020, we estimated an expected 2294 (95% CI 1872, 2716) cases of severe aortic stenosis who have not received treatment.

**Conclusion:** This study has demonstrated a significant decrease in TAVR and SAVR activity in England following the COVID-19 outbreak. This situation should be monitored closely, to ensure that monthly activity rapidly returns to expected levels. There is potential for significant backlog in the near-to-medium term, and potential for increased mortality in this population.

## Introduction

The pandemic of the novel coronavirus disease 2019 (COVID-19) [1] presents a global health crisis and has resulted in significant excess mortality worldwide [2]. As a result, many countries have imposed restrictions based on physical/social distancing and on movement (i.e. ‘lockdowns’), with the aim of mitigating and managing the spread of COVID-19. In the UK, the first confirmed case was recorded on 28th January 2020, with the first fatality from COVID-19 being announced on 5th March 2020 [3]. A UK-wide lockdown was initiated on 23rd March 2020.

The lockdown restrictions, and the pandemic itself, have resulted in widespread changes in operational activity of health services. Many countries and healthcare systems have faced significant pressure on services, particularly within critical care [4,5]. This has necessitated the need for restructuring of resources to meet those needs. Simultaneously, COVID-19 has influenced the ways in which people interact with health services. For example, previous studies have illustrated that there have been significant decreases in the number of admissions and diagnosis of many health conditions, including for acute coronary syndromes [6–10], stroke [11,12] and cancer referrals [13]. It is crucial to understand the consequences of this on public health and on future healthcare resource requirements, especially for conditions requiring timely healthcare interventions due to their adverse effect on prognosis, such as severe symtomatic aortic stenosis.

Aortic stenosis is the most common valvular heart disease and, while many patients with aortic stenosis are asymptomatic, the onset of symptoms is associated with rapid deterioration. Thus, timely treatment by either surgical aortic valve replacement (SAVR) or transcatheter aortic valve replacement (TAVR) is key. SAVR has been the default treatment strategy for symptomatic aortic stenosis, although TAVR has emerged as an effective option across operative-risk strata [14–18]. While the activity and outcomes for aortic valve replacements (surgical and transcatheter) have been studied in historic cohorts [19], there is a lack of data in contemporary practice, especially surrounding the impact of COVID-19 from a national perspective.

Therefore, the aim of this study was to investigate the activity and post-procedural outcomes of all aortic valve replacements (AVRs) in contemporary practice, from a national perspective, and to investigate the indirect impact of COVID-19 on activity and outcomes. The intention is to estimate the effect of reduced activity on projected backlog of cases.

## Methods

### UK TAVR Registry

The UK TAVR registry collects patient demographics, risk factors for intervention, procedural details, and in-hospital adverse outcomes/complications for every TAVR procedure undertaken within the UK [20]. Data collection occurs prospectively at each contributing centre, and is submitted to the National Institute for Cardiovascular Outcomes Research (NICOR). Data collection is mandated for all centres licensed to undertake TAVR procedures. We extracted data from NICOR on all TAVR procedures undertaken in England between January 2017, and June 2020.

### UK National Adult Cardiac Surgery Audit

The NICOR National Adult Cardiac Surgery Audit (NACSA) contains data on all major heart operations undertaken in the UK and Ireland [21]. We extracted all surgical aortic valve replacements (SAVRs) undertaken in England between 2017 and 2020. We defined SAVR to be any procedure that was recorded in NASCA as being a valve replacement procedure, and where the aortic valve implant type was recorded as being mechanical, biological, homograft or autograft replacement. Moreover, we further categorised SAVRs into (i) isolated AVR, (ii) AVR with coronary artery bypass graft (AVR+CABG), or (iii) AVR with other surgery (AVR+Other). Here, “other surgery” was defined as any mitral valve procedure, tricuspid valve procedure, pulmonary valve procedure, major aortic surgery, or other cardiothoracic procedures.

For both the UK TAVR registry and the NACSA registry, the British Cardiovascular Intervention Society and the Society for Cardiothoracic Surgery have made significant efforts to maintain data flows during this period, and have provided weekly uploads of data.

### Outcomes

The primary outcome considered in this study was presentation and treatment of severe symptomatic aortic stenosis with either SAVR or TAVR. As secondary outcomes, we considered 30-day mortality and post-procedural length of stay. Mortality information was provided by linking the UK TAVR registry and the NACSA with the office for national statistics civil registration of deaths dataset. Linkage was made based on each patient’s NHS number (a unique identifier given to all patients resident in England). Given that death registration is mandatory, this mortality data is definitive. We defined post-procedural length of stay to be the number of days between the TAVR/SAVR procedure and hospital discharge.

### Statistical Analysis

We excluded any cases in which the age of the patient at the time of the procedure was under 18 years. Additionally, we excluded cases where the NHS number was missing (since this variable was used for mortality linkage) or with missing procedure urgency (since this is a crucial variable in describing activity of TAVR/SAVR). Finally, we removed any duplicate cases in either datasets, with duplicates identified using NHS number, age at the time of operation, sex, admission date, and date/time of the procedure.

In all analyses, we stratified by procedure type (i.e. isolated AVR, AVR+CABG, AVR+Other or TAVR). Our aim was not to compare procedure types, but rather to explore features of each therapy individually. We made no formal comparisons between these procedural types, since there are a number of confounding factors surrounding the decision-making between TAVR/SAVR (many of which are not captured in the dataset) - that is, due to potential for confounding by treatment indication. Thus, all analyses focus on within-procedure-type comparisons, rather than between-procedure-type comparisons.

Cases performed between 1st February and June 2020 were defined into a “during COVID-19” group, with any case performed in these same months across the preceding years (i.e. 2017-2019) being defined into a “pre-COVID-19” group. The 1st February 2020 was chosen since the first COVID-19 case reported in England was on 28th January 2020. A sensitivity analysis was undertaken, where we took 1st April as the cut-off point (based on date of UK lockdown); results were quantitatively similar to those reported here (available on request).

We report patient baseline characteristics for each procedure type (i.e. isolated AVR, AVR+CABG, AVR+Other or TAVR), as whole cohorts and across the “during COVID-19” and “pre-COVID-19” groups. Continuous variables were reported using the mean with standard deviations. Categorical variables were presented as frequencies of occurrence with relative percentages. Temporal changes in predicted procedural risk are displayed by plotting the monthly average of the UK TAVR prediction model [22] for all TAVR procedures, and the Logistic EuroSCORE [23] for all SAVR procedures through time. These models were chosen for their respective uses as benchmarking tools for TAVR and SAVR. For the purposes of calculating the risk predictions, missing values in any predictor variables were set to “risk factor absent”, representing a plausible missingness process in the registries [19,22,24].

The number of procedures per month was calculated across the full study period, separately for isolated AVR, AVR+CABG, AVR+Other and TAVR. Percentage increase or decrease in monthly activity was calculated for each “during COVID-19” month, against the respective “pre-COVID-19” months. Similarly, we calculated the minimum, mean and maximum number of TAVR/SAVR procedures per month across 2017 and 2019, and compared these visually with the corresponding monthly counts in 2020 (up until June 2020). We fitted negative binomial models to the number of TAVR/SAVR procedures performed per month between January 2017 and December 2019, using time as covariates, which was modelled both continuously to capture increasing/decreasing trends in outcome and as a factor variable of month to capture any seasonality. This model was used to estimate the expected number of TAVR/SAVR procedures per month in 2020, to compare with the observed activity level.

For each of the four procedural types (i.e. isolated AVR, AVR+CABG, AVR+Other or TAVR), we compared mortality up to 30-days, across the “during COVID-19” and “pre-COVID-19” groups by fitting a Cox proportional hazards model, with the COVID-19 group as a covariate. Additionally, we adjusted for procedural risk by including the linear predictor of the UK TAVR prediction model [22] or the Logistic EuroSCORE [23], as appropriate. Differences in post-procedural length of stay between the “during COVID-19” and “pre-COVID-19” groups were also investigated by fitting a Cox proportional hazards model, with the aforementioned variables as covariates. The proportional hazards assumption was checked by examining the Schoenfeld residuals.

All analyses were performed using R version 4.0.0 [25], along with the tidyverse suite of packages [26], and the survival package [27,28].

## Results

The UK TAVR registry included 13995 procedures across the study period, of which we included 13376 in our analysis after applying the inclusion and exclusion criteria. The NACSA registry included 97108 cases over the study period, of which we included 26171 SAVR procedures, comprised of 12328 (47.1%) isolated AVR, 7829 (29.9%) AVR+CABG, and 6014 (23%) AVR+Other cases.

**Table 1** shows the baseline characteristics of the isolated AVR, AVR+CABG, AVR+Other and TAVR cases included in this study. The mean age of isolated AVR, AVR+CABG, AVR+Other and TAVR was 67.7, 72.3, 62.9 and 81.3, respectively. Across all surgical groups, the majority of cases were male. For the surgical AVR groups, the mean Logistic EuroSCORE was 7.52%, 10.7% and 13.9% for isolated AVR, AVR+CABG and

**Table 1:** *Baseline characteristics of the SAVR and TAVR cases included in this analysis. TAVR: Transcatheter Aortic Valve Replacement; SAVR: Surgical Aortic Valve Replacement. Note: the numbers in some categories might not sum to the total due to missing data*.

|  | Isolated<br>AVR | AVR+CABG | AVR+Other | TAVR |
| --- | --- | --- | --- | --- |
| n | 12328 | 7829 | 6014 | 13376 |
| Age, years (mean (SD)) | 67.74<br>(11.66) | 72.32 (8.20) | 62.87<br>(14.03) | 81.27<br>(7.27) |
| Female (%) | 4713<br>(38.2) | 1766 (22.6) | 2030 (33.8) | 5971<br>(44.7) |
| CCS Angina status (%) |  |  |  |  |
| No angina | 7056<br>(57.4) | 2336 (30.0) | 3999 (66.7) | 9467<br>(75.7) |
| Class I | 1497<br>(12.2) | 811 (10.4) | 657 (11.0) | 893 ( 7.1) |
| Class II | 2606<br>(21.2) | 2745 (35.2) | 847 (14.1) | 1493<br>(11.9) |
| Class III or IV | 1124 ( 9.2) | 1904 (24.4) | 496 ( 8.3) | 647 ( 5.2) |
| NYHA (%) |  |  |  |  |
| Class I | 1275<br>(10.5) | 795 (10.3) | 940 (15.9) | 860 ( 7.0) |
| Class II | 4854<br>(39.8) | 3203 (41.5) | 2052 (34.8) | 2851<br>(23.0) |
| Class III or IV | 6058<br>(49.7) | 3714 (48.2) | 2910 (49.3) | 8658<br>(70.0) |
| Previous MI (%) | 721 ( 5.9) | 1995 (25.6) | 290 ( 4.9) | 1916<br>(15.1) |
| Previous PCI (%) | 623 ( 5.1) | 911 (11.8) | 213 ( 3.6) | 2153<br>(17.0) |
| Previous Cardiac Surgery (%) | 846 ( 7.4) | 198 ( 2.7) | 848 (15.3) | 2150<br>(17.8) |
| Diabetic (%) | 2323<br>(18.9) | 2265 (29.0) | 717 (11.9) | 3057<br>(24.2) |
| Current/Ex Smoker (%) | 6469<br>(52.9) | 4805 (61.8) | 2991 (50.1) | 5819<br>(50.6) |
| Creatinine, umol/L (mean (SD)) | 89.51<br>(45.54) | 95.79 (53.59) | 96.28<br>(58.86) | 104.84<br>(61.84) |
| History of neurological disease (%) | 1059 ( 8.6) | 787 (10.1) | 592 ( 9.9) | 1992<br>(15.6) |
| Extracardiac Arteriopathy (%) | 623 ( 5.1) | 1020 (13.0) | 438 ( 7.3) | 1678<br>(13.4) |
| Atrial Fibrillation or Flutter (%) | 1109 ( 9.1) | 1056 (13.5) | 1405 (23.6) | 3385<br>(28.4) |
| One or more vessel with >50%<br>diameter stenosis (%) | 562 ( 5.1) | 6870 (96.9) | 236 ( 5.0) | 3659<br>(32.1) |
| PA Systolic (mean (SD)) | 27.55<br>(20.07) | 30.01 (23.30) | 39.38<br>(21.94) | 37.94<br>(15.15) |
| LV Function (%) |  |  |  |  |
| Good (LVEF > 50%) | 9739<br>(79.7) | 5512 (70.9) | 4239 (71.1) | 8913<br>(72.3) |
| Moderate (LVEF 31 - 50%) | 1918<br>(15.7) | 1792 (23.1) | 1375 (23.1) | 2281<br>(18.5) |
| Poor (LVEF < 30%) | 567 ( 4.6) | 467 ( 6.0) | 345 ( 5.8) | 1132 ( 9.2) |
| Height, m (mean (SD)) | 1.68 (0.10) | 1.70 (0.10) | 1.71 (0.11) | 1.65 (0.10) |
| Weight, kg (mean (SD)) | 82.73<br>(18.24) | 83.45 (16.83) | 82.36<br>(18.81) | 75.81<br>(17.30) |
| Non-Elective (%) | 2769<br>(22.5) | 2472 (31.6) | 2022 (33.6) | 2590<br>(19.4) |
| Logistic EuroSCORE (mean (SD)) | 7.52 (8.47) | 10.67 (11.07) | 13.93<br>(14.39) | NaN (NA) |
| UK TAVR CPM (mean (SD)) | NaN (NA) | NaN (NA) | NaN (NA) | 3.13 (2.43) |

AVR+Other, respectively, while the mean UK TAVR prediction model was 3.13% (Table 1).

### TAVR and SAVR Activity

There has been a steady increase in the number of TAVR procedures performed per month between January 2017 and December 2019, with the majority of procedures being elective (**Figure 1**). While the number of monthly AVR+CABG and AVR+Other procedures remained relatively stable pre-2020, there was a slight decrease in the number of elective isolated AVR cases per month in 2019. Specifically, the average number of elective isolated AVR cases per month was 250 in 2017 and 252 in 2018, while the monthly activity in 2019 decreased from 226 cases in January, to 151 cases by December (**Figure 1**). This trend continued into early 2020, with isolated AVR and AVR+CABG activity being lower throughout 2020, compared with historic levels (**Figure 2**). After 1st March 2020 there was a significant drop in activity across all AVR procedures compared with historic levels (**Figure 2**). There was a slight recovery in AVR activity in May and June 2020.

**Figure 1:**
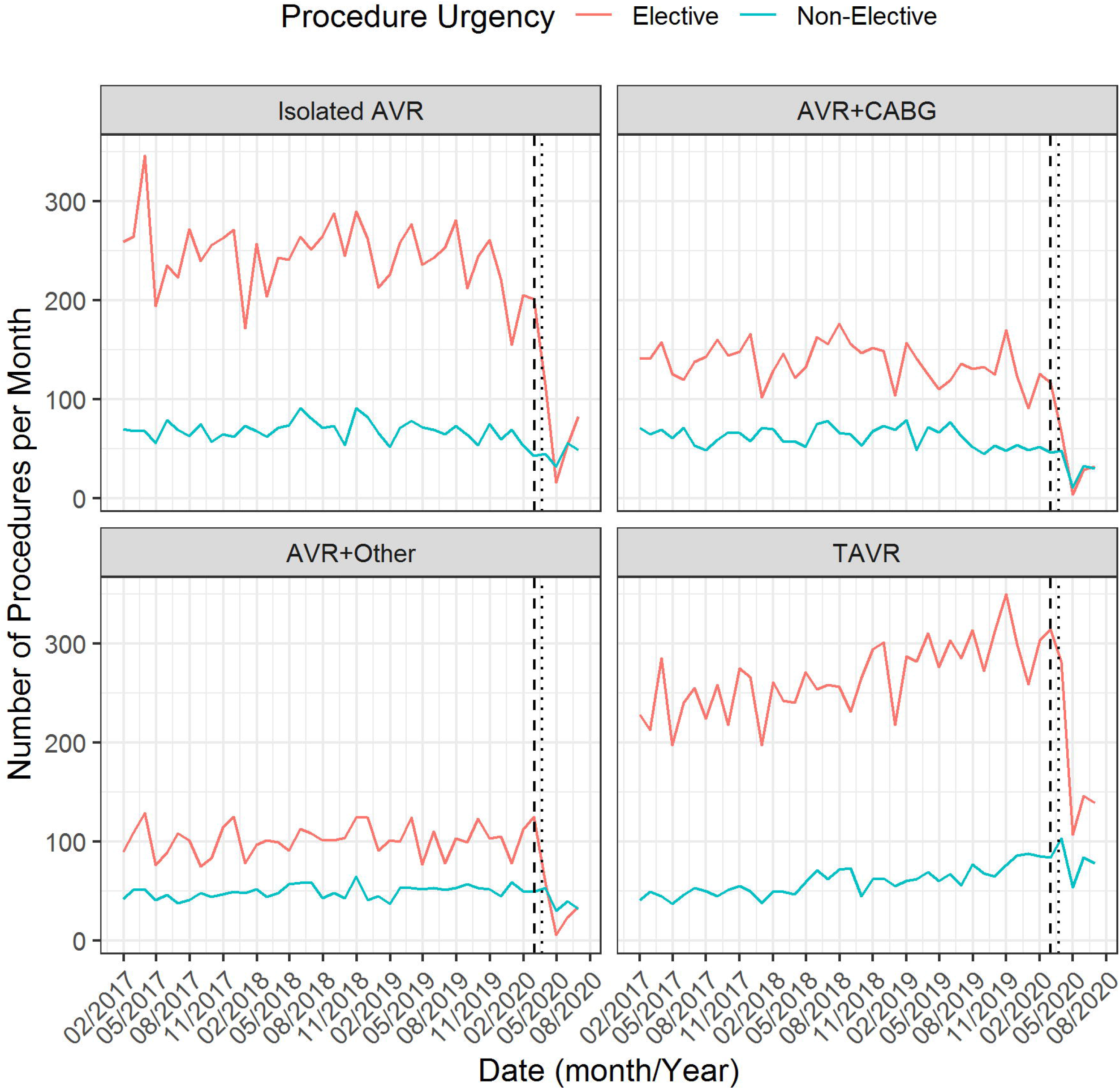
Temporal plot of the number of TAVR and SAVR procedures per month, stratified according to procedure urgency. The vertical dotted line denotes 23rd March 2020 (date of UK lockdown) with 1st March denoted by the vertical dashed line. TAVR: Transcatheter Aortic Valve Replacement; SAVR: Surgical Aortic Valve Replacement.

**Figure 2:**
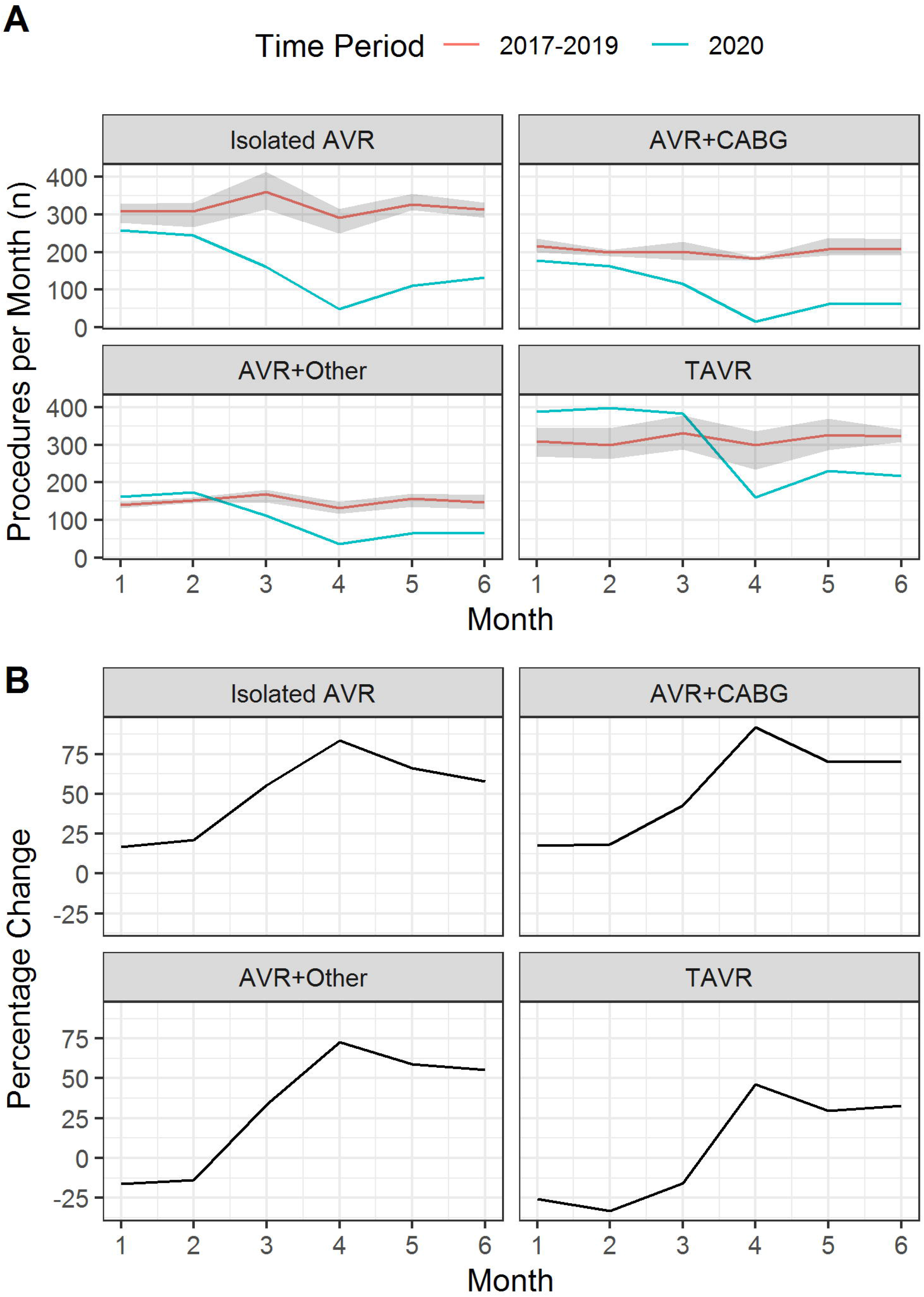
Panel A: Temporal plot of the number of TAVR and SAVR procedures per month during 2020, compared with monthly averages (minimum and maximum shown by shaded region) across all other years in the dataset. Panel B: Percentage change between the mean monthly activity in 2017-2019 and the monthly activity in 2020; negative percentage change denotes increase in 2020 over historic levels. TAVR: Transcatheter Aortic Valve Replacement; SAVR: Surgical Aortic Valve Replacement.

The drop in AVR activity was particularly pronounced for elective cases. The number of elective SAVR procedures, was below 100 cases per month after March 2020 for each of isolated AVR, AVR+CABG and AVR+Other (**Figure 1**). In contrast, while the number of elective TAVR cases also dropped after March 2020, there remained >100 elective TAVR cases per month. The relative drop (percentage change) in monthly activity between 2020 and historic levels was lower for TAVR than SAVR with a maximum percentage difference of 83.5%, 91.8%, 72.6% and 46.3%, for isolated AVR, AVR+CABG, AVR+Other and TAVR, respectively (**Figure 2, panel B**).

Fitting a negative binomial model to the monthly TAVR and SAVR procedures between 2017 and 2019 shows the expected and actual monthly AVR activity during 2020 (Figure 3). Specifically, the estimated difference (95% CI) in the number of TAVR cases per month compared with those expected based on historic trends was −41 (−79, −3) in March 2020, −222 (−257, −187) in April 2020, −188 (−226, −150) in May 2020, and −197 (−234, −159) in June 2020 (**Figure 3, panel B**). The estimated decrease in Isolated AVR activity was −189 (−220, − 158), −233 (−260, −207), −206 (−235, −177) and −171 (−199, −143), across March-June 2020, respectively. Similar observations were made for AVR+CABG and AVR+Other cases (**Figure 3, panel B**). Cumulatively, over the period March to June 2020, this amounts to an estimated expected drop of 2294 (95% CI 1872, 2716) cases of severe aortic stenosis in England, of which 799 (95% CI 685, 913) were for isolated AVR, 488 (95% CI 407, 570) were for AVR+CABG, 358 (95% CI 280, 436) were for AVR+Other, and 648 (95% CI 500, 796) were for TAVR.

**Figure 3:**
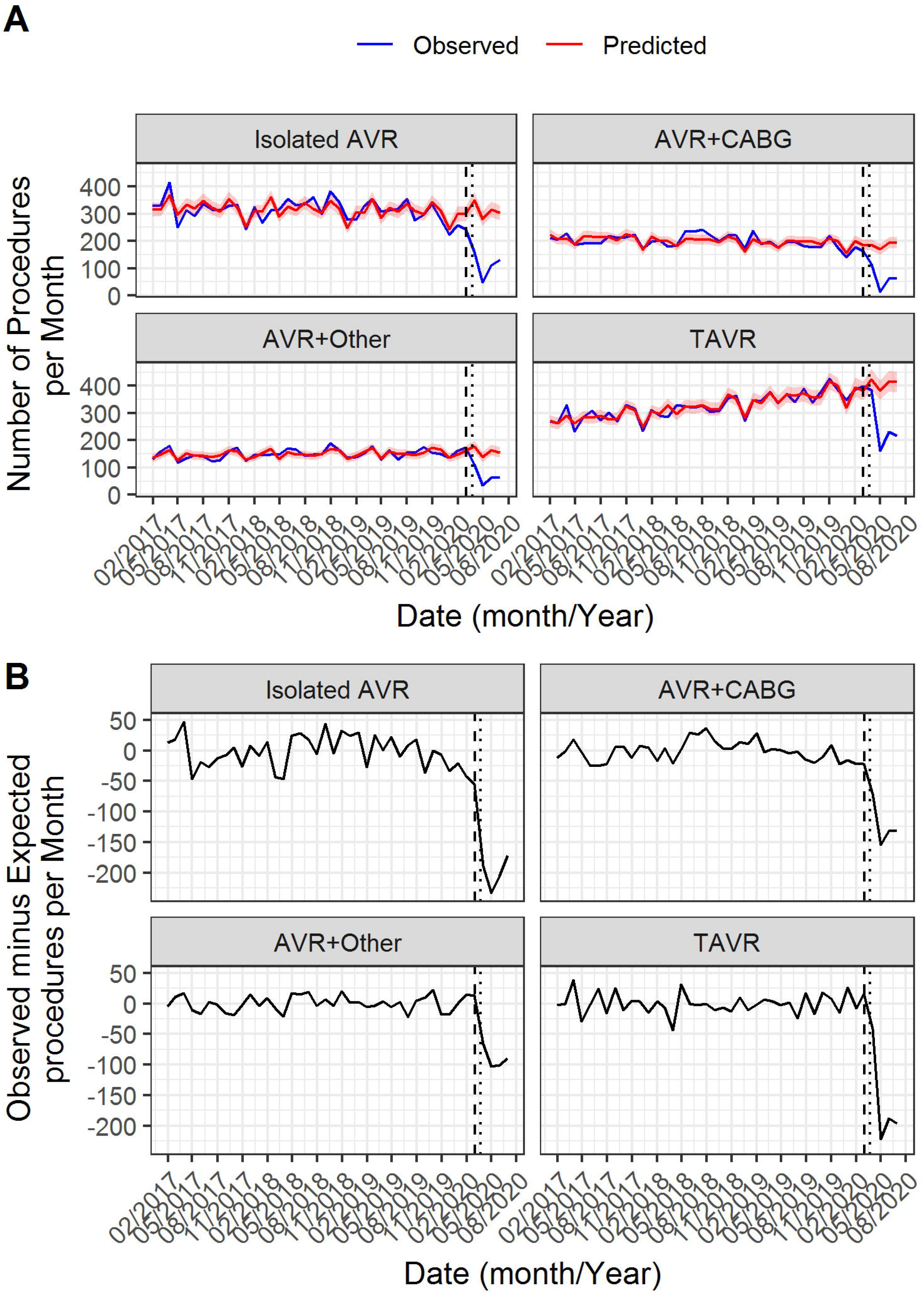
Panel A: Temporal plot of the observed and expected number of TAVR/SAVR procedures per month. Panel B: The difference between the observed and expected number of TAVR/SAVR procedures per month. In both plots the expected monthly count is estimated from a negative binomial model fitted to the 2017-2019 data. The vertical dotted line denotes 23rd March 2020 (date of UK lockdown) with 1st March denoted by the vertical dashed line. TAVR: Transcatheter Aortic Valve Replacement; SAVR: Surgical Aortic Valve Replacement.

### Evolution of Patient Demographics and Procedural Risk

**Table 2** shows patient baseline characteristics of isolated AVR cases across the pre-COVID-19 and during-COVID-19 groups. For isolated AVR, the mean age was significantly lower in the during-COVID-19 group than the pre-COVID-19 group (p<0.001), and there was a significantly higher CCS angina status (p<0.001), NYHA class (p<0.001) and mean PA systolic pressure (p<0.001). For AVR+CABG, there was no evidence of a significant difference in the majority of patient demographics between the during-COVID-19 group and the pre-COVID-19 group (**Supplementary Table 1**). Specifically, NYHA class (p<0.001) and mean PA Systolic pressure (p=0.004) were significantly higher in the during-COVID-19 group compared to the pre-COVID-19 group. For AVR+Other, there was a significantly higher proportion of male patients (p=0.001, **Supplementary Table 2**), and a higher NYHA class (p=0.029), between the during-COVID-19 group and the pre-COVID-19 group. For TAVR cases, the mean age, proportion of current/ex smokers and mean creatinine were significantly lower in the during-COVID-19 group compared with the pre-COVID-19 group (**Table 3**). Additionally, there was a lower proportion of TAVR cases with previous MI (p<0.001), previous cardiac surgery (p<0.001), and extracardiac arteriopathy (p=0.010) in the during-COVID-19 group, compared with the pre-COVID-19 group. The distribution of procedure urgency categories was significantly different between the during-COVID-19 group and the pre-COVID-19 group, for isolated AVR (p<0.001, **Table 2**), AVR+CABG (p=0.001, **Supplementary Table 1**), AVR+Other (p<0.001, **Supplementary Table 2**), and TAVR (p<0.001, **Table 3**).

**Table 2:** *Baseline characteristics of the Isolated AVR cases included in the pre-COVID-19 and during-COVID-19 groups, as defined in the methods section. AVR: Aortic Valve Replacement. Note: this only included cases in Febuary-June each year, and the numbers in some categories might not sum to the total due to missing data*.

|  | Pre-Covid-19 | During-Covid-19 | p-value |
| --- | --- | --- | --- |
| n | 4806 | 694 |  |
| Age, years (mean (SD)) | 67.87 (11.60) | 65.85 (10.84) | <0.001 |
| Female (%) | 1875 (39.0) | 249 (35.9) | 0.123 |
| CCS Angina status (%) |  |  | <0.001 |
| No angina | 2788 (58.3) | 372 (53.8) |  |
| Class I | 558 (11.7) | 86 (12.4) |  |
| Class II | 1013 (21.2) | 138 (20.0) |  |
| Class III or IV | 422 ( 8.8) | 95 (13.7) |  |
| NYHA (%) |  |  | <0.001 |
| Class I | 481 (10.1) | 48 ( 7.1) |  |
| Class II | 1895 (39.9) | 230 (33.8) |  |
| Class III or IV | 2377 (50.0) | 402 (59.1) |  |
| Previous MI (%) | 275 ( 5.7) | 32 ( 4.6) | 0.269 |
| Previous PCI (%) | 214 ( 4.5) | 39 ( 5.6) | 0.227 |
| Previous Cardiac Surgery (%) | 347 ( 7.8) | 46 ( 7.2) | 0.620 |
| Diabetic (%) | 931 (19.5) | 142 (20.5) | 0.567 |
| Current/Ex-Smoker (%) | 2521 (52.9) | 370 (53.9) | 0.666 |
| Creatinine, umol/L (mean (SD)) | 89.36 (43.83) | 87.29 (42.29) | 0.254 |
| History of neurological disease (%) | 390 ( 8.2) | 60 ( 8.7) | 0.666 |
| Extracardiac Arteriopathy (%) | 242 ( 5.0) | 29 ( 4.2) | 0.379 |
| Atrial Fibrillation or Flutter (%) | 449 ( 9.4) | 49 ( 7.1) | 0.060 |
| One or more vessel with >50% diameter stenosis (%) | 223 ( 5.2) | 35 ( 5.7) | 0.732 |
| PA Systolic (mean (SD)) | 26.00 (19.88) | 36.57 (31.66) | <0.001 |
| LV Function (%) |  |  | 0.456 |
| Good (LVEF > 50%) | 3778 (79.4) | 534 (77.4) |  |
| Moderate (LVEF 31 - 50%) | 758 (15.9) | 119 (17.2) |  |
| Poor (LVEF < 30%) | 222 ( 4.7) | 37 ( 5.4) |  |
| Height, m (mean (SD)) | 1.68 (0.10) | 1.69 (0.11) | 0.041 |
| Weight, kg (mean (SD)) | 82.77 (18.18) | 84.56 (18.97) | 0.016 |
| Non-Elective (%) | 1074 (22.3) | 225 (32.4) | <0.001 |
| Logistic EuroSCORE (mean (SD)) | 7.48 (8.23) | 6.90 (8.17) | 0.080 |

**Table 3:** *Baseline characteristics of the TAVR cases included in pre-COVID-19 and during-COVID-19 groups, as defined in the methods section. TAVR.: Transcatheter Aortic Valve Replacement. Note: this only included cases in Febuary-June each year, and the numbers in some categories might not sum to the total due to missing data*.

|  | Pre-Covid-19 | During-Covid-19 | p-value |
| --- | --- | --- | --- |
| n | 4743 | 1390 |  |
| Age, years (mean (SD)) | 81.36 (7.36) | 80.65 (7.12) | 0.001 |
| Female (%) | 2164 (45.7) | 615 (44.2) | 0.363 |
| CCS Angina status (%) |  |  | 0.071 |
| No angina | 3376 (75.1) | 981 (78.0) |  |
| Class I | 344 ( 7.6) | 72 ( 5.7) |  |
| Class II | 537 (11.9) | 146 (11.6) |  |
| Class III or IV | 240 ( 5.3) | 59 ( 4.7) |  |
| NYHA (%) |  |  | 0.072 |
| Class I | 306 ( 6.9) | 84 ( 6.7) |  |
| Class II | 1015 (22.7) | 250 (19.8) |  |
| Class III or IV | 3141 (70.4) | 928 (73.5) |  |
| Previous MI (%) | 746 (16.3) | 146 (11.4) | <0.001 |
| Previous PCI (%) | 770 (17.0) | 190 (14.8) | 0.066 |
| Previous Cardiac Surgery (%) | 826 (19.0) | 178 (13.9) | <0.001 |
| Diabetic (%) | 1090 (23.9) | 312 (24.4) | 0.756 |
| Current/Ex-Smoker (%) | 2058 (48.9) | 508 (45.4) | 0.041 |
| Creatinine, umol/L (mean (SD)) | 105.73 (66.89) | 101.14 (56.44) | 0.029 |
| History of neurological disease (%) | 718 (15.6) | 205 (15.9) | 0.786 |
| Extracardiac Arteriopathy (%) | 641 (14.2) | 143 (11.3) | 0.010 |
| Atrial Fibrillation or Flutter (%) | 1253 (28.9) | 353 (28.3) | 0.705 |
| One or more vessel with >50% diameter stenosis (%) | 1313 (31.6) | 332 (29.2) | 0.131 |
| PA Systolic (mean (SD)) | 38.22 (15.02) | 37.12 (15.60) | 0.139 |
| LV Function (%) |  |  | 0.162 |
| Good (LVEF > 50%) | 3224 (72.4) | 877 (71.9) |  |
| Moderate (LVEF 31 - 50%) | 836 (18.8) | 214 (17.6) |  |
| Poor (LVEF < 30%) | 394 ( 8.8) | 128 (10.5) |  |
| Height, m (mean (SD)) | 1.65 (0.10) | 1.66 (0.10) | 0.055 |
| Weight, kg (mean (SD)) | 75.66 (17.37) | 76.51 (17.47) | 0.126 |
| Non-Elective (%) | 832 (17.5) | 403 (29.0) | <0.001 |
| UK TAVR CPM (mean (SD)) | 3.23 (2.76) | 2.89 (1.80) | <0.001 |

Overall procedural risk, as estimated by the Logistic EuroSCORE, has remained relatively stable over time for isolated AVR, AVR+CABG and AVR+Other (**Supplementary Figure 1**). There was no significant difference in the mean Logistic EuroSCORE between the pre-COVID-19 groups and during-COVID-19 groups for isolated AVR, AVR+CABG or AVR+Other cases (**Table 2**, **Supplementary Table 1** and **Supplementary Table 2**). For TAVR, overall procedural risk, as estimated by the UK TAVR prediction model, dropped significantly over 2017, with this stabilizing to approximately 4% for non-elective cases and approximately 2.5% for elective cases (**Supplementary Figure 1**). While the mean UK TAVR prediction model was significantly lower in the during-COVID-19 group compared with the pre-COVID-19 group (p<0.001, Table 3), this was largely driven by 2017 cases (**Supplementary Figure 1**).

Between 2017 and December 2019, there has been a steady increase in the monthly TAVR activity in the lowest quantiles of risk strata (estimated by UK TAVR prediction model), while the monthly activity in the highest quantiles of risk strata as remained relatively stable (**Supplementary Figure 2**). After March 2020, TAVR activity was reduced across all quantiles of risk strata. In contrast, the monthly activity of isolated AVR, AVR+CABG and AVR+Other has been gradually decreasing through time across all quantiles of risk strata (defined by Logistic EuroSCORE), with this continuing post March 2020 (**Supplementary Figure 3, Supplementary Figure 4**, and **Supplementary Figure 5**).

### Outcomes

The overall (unadjusted) Kaplan-Meier estimates of 30-day survival were 98.5% for isolated AVR, 95.9% for AVR+CABG, 94.9%, for AVR+Other, and 97.5% for TAVR. We found no significant difference in mortality hazards up to 30-days post procedure between the pre-COVID-19 group and the during-COVID-19 group for isolated AVR, AVR+CABG, AVR+Other or TAVR (Table 4).

**Table 4:** *Multivariable adjusted mortality hazard ratios (95% confidence interval) of COVID-19 period (during-vs. pre-) for up-to 30-day mortality. All values are adjusted for overall procedural risk (Logistic EuroSCORE for SAVR and UK TAVR prediction model for TAVR cases). TAVR.: Transcatheter Aortic Valve Replacement; SAVR: Surgical Aortic Valve Replacement*.

| Surgical Group | Hazard Ratio (95% CI) | p-value |
| --- | --- | --- |
| Isolated AVR | 1.15 (0.63, 2.12) | 0.643 |
| AVR+CABG | 1.19 (0.77, 1.83) | 0.431 |
| AVR+Other | 1.01 (0.63, 1.61) | 0.983 |
| TAVR | 0.7 (0.45, 1.07) | 0.098 |

The median length of stay (LOS) following TAVR was 3 days (interquartile range: 2-5 days) in the pre-COVID-19 group, and 2 days (interquartile range: 1-3 days) in the during-COVID-18 group. The median (interquartile range) LOS in the pre-COVID-19 group for isolated AVR, AVR+CABG and AVR+Other was 7 (5-9) days, 8 (6-12) days and 9 (6-15) days, respectively, with these being 6 (5-9) days, 7 (5-10) days and 8 (6-13) days in the during-COVID-19 group. For AVR+CABG and AVR+Other procedures performed in the during-COVID-19 period, the adjusted hazard ratios (95% CI) for early discharge were 1.21 (1.08, 1.35)and 1.16 (1.04, 1.3), respectively, showing significantly shorter LOS (Table 5). For TAVR, the hazard ratio for shorter LOS was modelled as a function of time to account for non-proportional hazards, with time demonstrating significantly shorter LOS for the during-COVID-19 group, up to 2 days post procedure.

**Table 5:** *Multivariable adjusted hazard ratios (95% confidence interval) for discharge across COVID-19 period (during-vs. pre-). The event is discharge, so a hazard ratio greater than one implies shorter length of stay. All values are adjusted for overall procedural risk (Logistic EuroSCORE for SAVR and UK TAVR prediction model for TAVR cases). TAVR: Transcatheter Aortic Valve Replacement; SAVR: Surgical Aortic Valve Replacement*.

| Surgical Group (period, where applicable) | Hazard Ratio (95% CI) | p-value |
| --- | --- | --- |
| Isolated AVR | 1.02 (0.94, 1.11) | 0.565 |
| AVR+CABG | 1.21 (1.08, 1.35) | p<0.001 |
| AVR+Other | 1.16 (1.04, 1.3) | 0.006 |
| TAVR |  |  |
| 0-1 Days | 2.87 (2.54, 3.24) | p<0.001 |
| 1-2 Days | 1.51 (1.34, 1.69) | p<0.001 |
| 2+ Days | 1.07 (0.95, 1.21) | 0.247 |

## Discussion

This national study is the first to investigate activity and outcomes following all aortic valve replacement procedures in contemporary practice, including the potential indirect impacts of the COVID-19 pandemic. We observed a rapid and significant decrease in TAVR and SAVR activity during the COVID-19 pandemic. We estimated that over the period March to June 2020, this decline in activity accounts for an estimated 2294 patients with severe aortic stenosis left untreated by TAVR or surgical intervention. This will have major implications on this cohort of patients whose untreated mortality is high. Nevertheless, we also report that despite restructure of healthcare services nationally during COVID-19, overall procedural risk has maintained relatively constant, and there was no evidence of a difference in procedure-related mortality outcomes during this period.

The treatment of severe aortic stenosis has evolved rapidly over the last decade, moving from SAVR being the default treatment modality, to a situation in which TAVR is now an evidence-based option at all surgical risk categories [14–18]. This study observed changes in TAVR and SAVR activity, with steadily increasing TAVR activity and corresponding slight decreases in elective isolated AVR cases, up-to 2019. This supports previous findings in this area [19]. Although TAVR is currently only approved for inoperable or high-risk cases in the UK, the evidence of equivalence in low-risk cases in accumulating [18,29]. This may partially explain our finding of a steadily increasing proportion of TAVR cases within the lowest quantile of risk (as quantified by the UK TAVR prediction model), prior to 2020. The observed decrease in SAVR activity (particularly elective isolated AVR cases) before COVID-19, also provides evidence that the clinical envelope for TAVR has expanded into lower risk cases within real-world contemporary practice.

Inevitably, the initiation of national lockdown in the UK was associated with a significant reduction in the monthly number of TAVR and SAVR procedures being performed. We found a maximum percentage decrease in monthly activity between January and June 2020 compared with corresponding historic levels of 83.5%, 91.8%, 72.6% and 46.3%, for isolated AVR, AVR+CABG, AVR+Other and TAVR, respectively, demonstrating a relatively smaller fall in TAVR than SAVR. Even between March and June 2020, there remained over 100 elective TAVR cases per month, compared with a near-complete reduction in elective SAVR. One potential explanation is that TAVR has a very low probability of requiring stay in an intensive treatment unit after the procedure, which is important given constraints during the COVID-19 pandemic. Indeed, during the pandemic, patients with severe symptomatic aortic stenosis were recommended to be treated by TAVR where appropriate [30]. However, given that the UK TAVR registry and the NACSA dataset does not contain information on the decision-making behind the SAVR or TAVR choice, we are not able to investigate this directly within this study.

The observed reduction in TAVR/SAVR activity was largely driven by elective cases, with this study showing that non-elective cases have remained relatively stable post March 2020. A possible explanation for this was that the UK government response to the COVID-19 pandemic was to recommend cancellation of elective procedures [31]. This was made to allow a re-structuring of hospital services, thereby allowing more staff and resource to deal with the increased admissions due to COVID-19. Simultaneously, this reduces exposure of individual patients and their relatives to the hospital environment, and reduces exposure of healthcare workers to asymptomatic COVID-19 patients. We observed that monthly activity for TAVR and SAVR had a slight recovery in May and June 2020, following the initial drop during March and April 2020. We were unable to investigate the reasons for this, but a similar observation was made for admissions for acute coronary syndrome [6]. Again, one could speculate that this relates to decreasing demands on healthcare systems as the pandemic evolved, with an aim to resume elective activity once the peak of the pandemic had passed.

While the observed temporal changes in TAVR and SAVR activity are perhaps unsurprising, the marked drop in cases post COVID-19 does raise important implications for healthcare resource planning in the near-to-medium term. Our estimates surrounding the difference in the number of AVR procedures per month in 2020 compared with historic trends mean that there will likely be significant increased demand for TAVR and SAVR. This will lead to an inevitable increase in waiting times [32], and associated adverse impacts on outcomes [33]. Recommendations for how to manage this challenge are emerging [32,34]. Since the NACSA and UK TAVR registry datasets only contain data on those who undergo TAVR/SAVR, it was not possible for us to forecast future demand for AVR (since we do not have information on patients who are candidates for AVR, but who have not currently undergone the procedure). However, based on the available data, we estimated that, cumulatively, between March and June 2020, there were 2294 (95% CI 1872, 2716) cases of severe aortic stenosis who have not received treatment in England. Previous studies have shown that, under normal circumstances, the median wait-time for TAVR is 80 days [35]. Thus, assuming these figures apply to AVR generally, one could postulate that this means approximately 1147 patients will remain untreated by 80 days (increasing to 1720 if procedures in the near-to-medium term are made at 50% capacity), even without considering the additional cases over this period. While such figures are an approximation, they do give some indication of expected levels of increased demand that SAVR and TAVR centres might expect as the lockdown restriction continues to be relaxed. On a national-level, it is clear that strategies will be required to mitigate this large backlog of cases, in order to reduce avoidable deaths in patients with severe symptomatic aortic stenosis who remain untreated.

Indeed, it remains unclear what effect the reduction in the number of procedures per month has had on outcomes for patients with aortic stenosis who would otherwise have been treated with AVR during March - June 2020. Some of these patients might be alive and waiting for AVR intervention, or might have died from aortic stenosis, COVID-19 or from other causes. Previous studies have estimated that the risk of death whilst waiting for intervention for server aortic stenosis in routine clinical practice is between 2% and 14% [36]. This means that of the estimated 2294 currently delayed cases, there will be between 46 and 321 deaths while waiting for intervention, if capacity returns to normal. Any additional delays due to the backlog will lead to increased mortality. Of course, these are approximate figures and does not account for excess mortality due to COVID-19 [37].

Several limitations should be noted when interpreting the results of this study. Firstly, we make no statistical comparisons between isolated AVR, AVR+CABG, AVR+Other or TAVR groups. Any such comparisons would be subject to confounding by indication. This means that we were not able to investigate changes in patient-level propensity to undergo SAVR vs. TAVR through the COVID-19 period, since the decision-making behind the SAVR vs. TAVR choice was not recorded in these data. Secondly, while we used the Logistic EuroSCORE to summarise overall SAVR procedural risk, this model is known to overpredict mortality risk. However, this model is commonly used for benchmarking in national cardiovascular registries, and we use the model in the same capacity here. Thirdly, this analysis is limited to procedures in England; however, given that COVID-19 has caused changes in healthcare utilisation globally, one might expect similar findings in other healthcare settings. Finally, some delays in reporting during the pandemic might contribute to some of the results; however, the British Cardiovascular Intervention Society and the Society for Cardiothoracic Surgery have made significant efforts to maintain data flows during this period, and have provided weekly uploads of data. Thus, this limitation is potentially minimised.

In conclusion, this study has demonstrated a significant drop in TAVR and SAVR activity following the COVID-19 outbreak in the UK. The case-mix of patients who have undergone SAVR or TAVR during the COVID-19 period was similar to the case-mix seen in the pre-COVID-19 period, with no evidence of a difference in mortality outcomes. The activity across both SAVR and TAVR should be closely monitored to ensure that monthly cases correctly return to expected levels. These data suggest that there will be a sharp rise in demand for AVR intervention in the near-to-medium term, with the potential for an upturn in mortality in patients waiting to be treated.

## Data Availability

Data will be available upon ethical approval.

## Acknowledgments

The authors acknowledge Chris Roebuck, Tom Denwood, Tony Burton and Courtney Stephenson and data support staff at the NHS digital for providing and creating the secure environment for data hosting and for analytical support, and Anil Gunesh and Julian Hains from the National Institute of Cardiovascular Outcomes Research for data transfer into the secure environment.

